# Updating Herd Immunity Models for the U.S. in 2020: Implications for the COVID-19 Response

**DOI:** 10.1101/2020.10.05.20207100

**Authors:** Natalie E. Sheils, Gregory D. Lyng, Ethan M. Berke

## Abstract

**Objectives:** To understand what levels of herd immunity are required in the COVID-19 pandemic, given spatial population heterogeneity, to best inform policy and action.

**Methods:** Using a network of counties in the United States connected by transit data we considered a set of coupled differential equations for susceptible-infectious-removed populations. We calculated the classical herd immunity level plus a version reflecting the heterogeneity of connections in the network by running the model forward in time until the epidemic completed.

**Results:** Necessary levels of herd immunity vary greatly from county to county. A population weighted average for the United States is 47.5% compared to a classically estimated level of 77.1%.

**Conclusions:** Common thinking argues that the nation needs to achieve at least 60% herd immunity to emerge from the COVID-19 pandemic. Heterogeneity in contact structure and individual variation in infectivity, susceptibility, and resistance are key factors that reduce the disease-induced herd immunity levels to 34.2–47.5% in our models. Looking forward toward vaccination strategies, these results suggest we should consider not just who is vaccinated but where those vaccinations will do the most good.

## Introduction

Classical infectious disease models assume that all people transmit and get infected with SARS-CoV-2 at the same rate (i.e., the population mixes homogeneously), but in reality, populations are heterogeneous. This is because most people interact only within a small circle of close contacts. Groups of teenagers tend to not interact with groups of people in their 60s. People in Montana do not tend to interact with people in New York. Only a few people move across and between groups. The standard calculation, based on classical epidemiological models for infectious disease, identifies a threshold for *herd immunity*. Assuming lasting immunity post infection and simple disease dynamics, this calculation shows that, once a sufficiently large fraction of the population has been infected/removed, the remaining susceptible population is too small to support a continuing epidemic. While transmission will still occur, new infections do not generate enough subsequent infections to perpetuate the outbreak. Thus, the herd immunity threshold represents a critical value that quantifies the level of immunity in a population that will prevent future major outbreaks.

The herd immunity threshold may be expressed in terms of the well-known reproduction number *R*_0_ as 1 − 1/*R*_0_. For example, the 1918 influenza pandemic is believed to have had an *R*_0_ of around 2.9 (1); this translates to a herd immunity threshold around 66%. Similarly, frequently cited and discussed estimates of herd immunity for COVID-19 requiring 60% or more of the population to be infected (2–8) are based on the same reasoning and an estimated *R*_0_ ≈ 2.5 (9). However, reconstructions of the 1918 pandemic suggest that only a third of the world’s population was actually infected during the course of the pandemic (10). Similarly, the H1N1 virus that set off the 2009 influenza pandemic had an estimated *R*_0_ between 1.2 and 1.6 (11); a value of 1.4 generates a classical herd immunity threshold of 28.5%. Yet, CDC estimates indicate that only 19.8% of the US population was infected (12). The classical formula is based on a modeling assumption of population homogeneity, and this is a key driver of the overestimates noted above.

There is now a robust scientific discussion (4,13–17) around quantifying the impact of these heterogeneities on herd immunity during the current pandemic. Our work advances this discussion by modeling the impact of spatial heterogeneity on herd immunity; our calculations address an issue identified as important by Britton et al. (13), namely, the impact of spatial heterogeneity, and, in particular, the role of rural areas with lower contact rates. We also incorporate a nonuniform immunity structure into the model, motivated by the presence, in some individuals, of helper T-cells that are cross-reactive to SARS-CoV-2. We demonstrate that the proportion of the population infected to achieve herd immunity may be lower than usually assumed, which would have significant implications for public health.

### Understanding immunity

A substantial proportion of individuals have pre-existing SARS-CoV-2 cross-reactive T cells due to repeated exposure to the four common cold coronaviruses (18,19). Because we are re-exposed annually to these viruses, an estimated 90% of adults have had coronavirus antibodies at some point, although protective antibodies from them only last between a few months to 34 years. Current research estimates that 35% of people who have never been exposed to SARS-CoV-2 have these helper T-cells due to exposure to these common coronaviruses (20). While the impact of these cells on disease progression and clinical outcomes remains to be fully explored, there are hints that they may contribute to partial immunity to the virus that causes COVID-19. Some suggest that some individuals may have an innate resistance to the disease (21). Our calculations show that these factors, representing another form of population heterogeneity, also impact the herd immunity threshold.

Building on these insights, Lourenço et al. (14) considered a simple two-compartment model in which one of the groups is resistant to infection (either by innate resistance or from cross-reactive protection gained by exposure to other seasonal coronaviruses). The results show that, depending on the magnitude of the resistant fraction, the value of the reproduction number, and the amount of mixing between groups, the computed level of herd immunity can vary dramatically. Given the uncertainty around the disease, the authors make the case that herd immunity may well have already been achieved in some locales. We incorporate a weakened version of this protection (representing the possibility of partial immunity) into our spatial model for disease progression.

### Spatial models - moving beyond homogeneous models of herd immunity

Britton et al. (13) made an early and important contribution to the herd immunity discussion for COVID-19 by presenting an age-structured model containing six age groups in a single community. In their example, the contact structure between age groups was based on an empirical formulation. The final calculation demonstrates clearly the principle that heterogeneous contact can lower herd immunity thresholds. For example, one numerical experiment suggests a herd immunity level around 43% (compared to a classical estimate of 60%).

Because it is the inspiration for our calculation, we describe their calculation in some detail. For each of the different population structures under consideration, the authors implement uniform (multiplicative) non-pharmaceutical interventions. In terms of the model, they simply multiply the next-generation matrix by a constant, positive factor a *α* < 1. This roughly simulates stay-at-home or other preventive measures. They then run the epidemic to its conclusion and expose the population to a second wave, but this time with *α* = 1; no restrictions. They define a *α*_∗_ to be the largest value of *α* for which there is no second epidemic. (One can think of it this way: In the heterogenous population, *α*_∗_ represents the “perfect” amount of social distancing; it represents the minimum amount of preventative measures required to achieve herd immunity upon the conclusion of the outbreak.) Finally, the disease-induced herd immunity threshold is computed as the proportion of the population that is infected in the first wave. To compute this requires a calculation of the time-asymptotic limiting state of the system.

## METHODS

The mathematical model is a large, coupled system of ordinary differential equations of SIR type. That is, the model utilizes compartmental structure of classical epidemiological models and features susceptible, infectious, recovered, and deceased compartments at each node in a geographical network. In this case, the nodes of the network represent counties (or county equivalents) in the continental US; see Figure 1. For these calculations, there are 1,829 counties in the network, so the system nominally consists of 7,316 ordinary differential equations. (Due to data availability and calibration considerations, not all counties in the continental US are represented; the counties included in the network represent 93% of the US population). Moreover, we connect the nodes with edges drawn based on transit data. In particular, we insert “short-range” edges between nodes using thresholds of movements based on publicly available commuter data while “long-range” edges are based on passenger movements on domestic airline flight patterns. The network thus creates a coarse, but realistic, representation of US geography and typical human movements between counties. Finally, we use these edges to weakly couple neighboring nodes. Parameters for county-level contact rates and the long-range and short-range coupling coefficients are estimated from March 2020 COVID-19 mortality data. We then compute *R*_0_ in standard fashion as the largest eigenvalue of the next generation matrix. From this value, we compute the classical herd immunity threshold for the model. We take conservative values for the relative recovery and disease-induced mortality rates (assumed to be the same at each node). Coupled with the estimated contact rates from early data, this results in rather high estimates for the reproduction number (and thus rather high classical estimates for herd immunity). Even so, our calculations suggest an actual disease-induced herd immunity level substantially below the ≥60% levels frequently cited for COVID-19. See the supplemental information for further details about the model and its parametrization.

**Figure 1:**
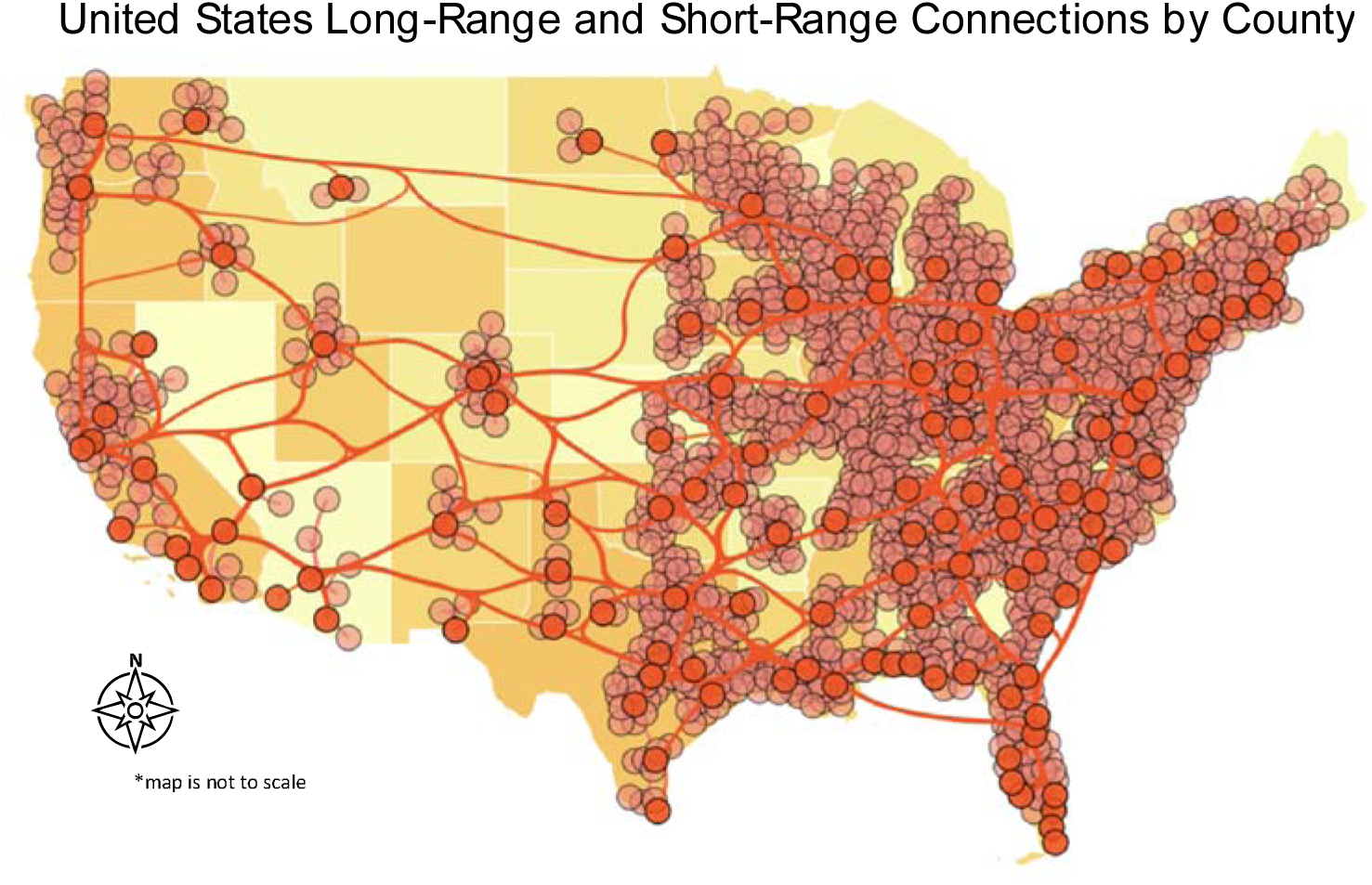
The network underlying the model. See the supplemental information for further details about the construction of the network.

To calculate the herd immunity threshold for this model, we seed each node with a small fraction of infectious people and run the simulations forward as described above. To incorporate partial immunity, we introduce a second infectious compartment at each node to account for those who are infectious but have cross-reactive T cells. In this version of the model, this fraction of individuals is assumed to be less infectious and to have, on average, a quicker recovery time.

## RESULTS

Calculations from the model featuring spatial heterogeneity predict herd immunity will be reached at around 47.5% (versus an 77.1% estimate based on the classical techniques and the calculation of *R*_0_ for this parametrization of the model). Figure 2 shows the variation in the proportion of each county that must be immune based on the disease-induced herd immunity calculation. The figure shows that, far from the idealized scenario implicit in classical epidemiological models, where immunity is assumed to be uniformly spread across the population, we should expect substantial variation across geographies in the required local immunity levels. We note that, qualitatively, this finding mirrors recent results from 1.46 million antibody tests in New York City, a one-time COVID-19 epicenter. The antibody tests suggest an overall prevalence of 27% with values in various ZIP codes ranging from 12.6% to 51.6% (22).

**Figure 2:**
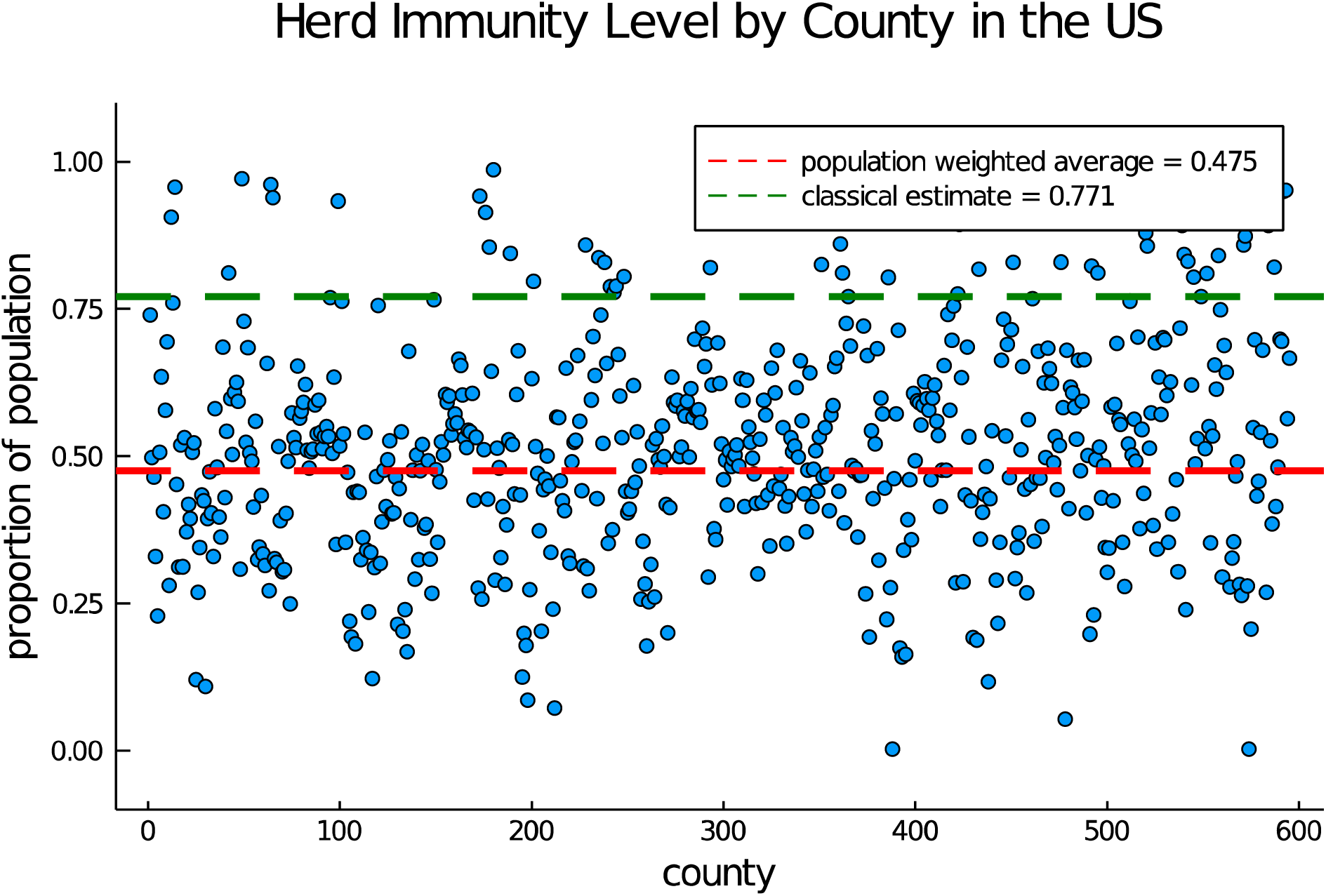
Spatial heterogeneity and herd immunity. Each dot represents the proportion of the population that must become “immune” in a given county in order to halt the spread of the disease. The classical estimate is found using the formula 1-1/*R*_0_ while our estimate is found via the population-weighted average of the level found at each county on a geographically heterogenous network. Only counties with more than 33 COVID-19 deaths as of September 14, 2020 as reported by the New York Times (29) are shown.

A similar calculation, this time assuming 35% of the population has cross-reactive T cells (20) and that they, on average, recover 1 day more quickly and that their disease-induced relative mortality rate is 10% less than those without cross-reactive cells, reduces the disease-induced estimate of herd immunity to 34.2%; see Figure 3. This calculation shows that even modest heterogeneity in infectiousness and duration of the disease course impacts the levels of disease-induced herd immunity.

**Figure 3:**
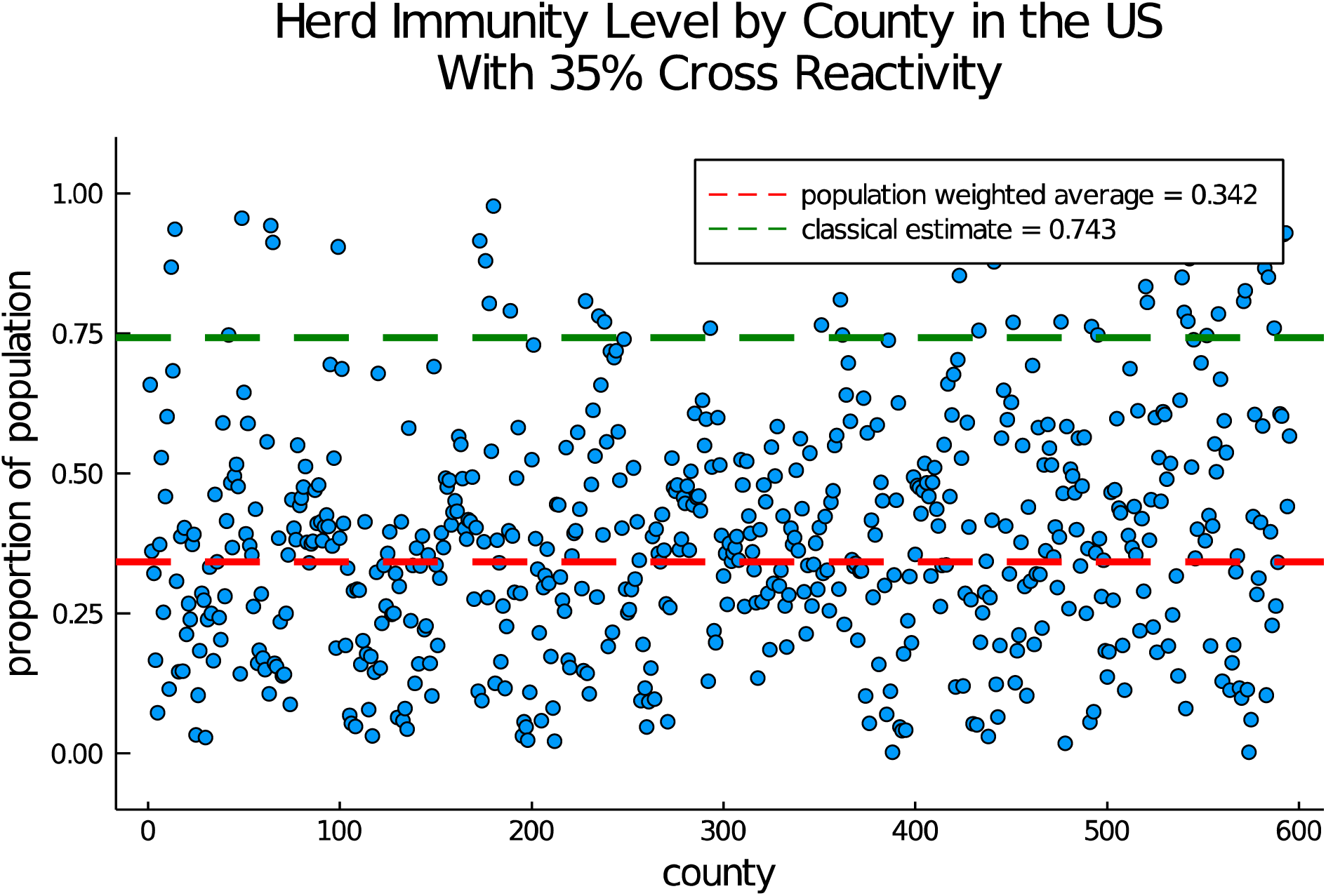
Herd immunity with cross reactivity. Each dot represents the proportion of the population needed to reach herd immunity in a single county. The classical estimate is found using the formula 1-1/*R*_0_ while our estimate is found via the population-weighted average of the level found at each county using the approach of Britton et al. (13) on a geographically heterogeneous network. Only counties with more than 33 COVID-19 deaths as of September 14, 2020 are shown.

## DISCUSSION

This work demonstrates that herd immunity may be achieved at a level significantly below the classical estimate of 60%, based on spatial modeling that better aligns to population distribution. The classical models’ assumption of a homogenously mixing population in which each individual is equally susceptible and equally infectious (once infected) and equally likely to contact each other individual is a gross oversimplification. Nonetheless, it is represented in much of the ongoing public discourse and public health messaging around the disease (2,3). In reality, heterogeneity manifests itself across multiple dimensions that are meaningful for disease transmission (e.g., in contact networks, in susceptibility, and in infectiousness). Thus, we should expect that the disease will exploit these heterogeneities so that an initial wave of infections will preferentially target especially vulnerable or highly connected individuals and that the conferral of immunity on (or removal of) these central-to-transmission individuals will stymy any “second wave” in the connected population.

We are now more than 8 months into the COVID-19 pandemic, and there is still much that we do not know. In particular, our understanding of immunity still has gaps. Much of the literature and public reporting has relied almost solely on antibody testing to tell the immunity story. The results of numerous seroprevalence studies have been published, and virtually none of them approach the 60% level. Among the remarkable examples are two former epicenters of the pandemic; see Table 1. On the one hand, these results have been used to indicate how “far” we are from herd immunity. For example, antibody testing results showing similar prevalence (17% in Stockholm in April 2020) and a reliance on classical estimates of herd immunity make up the key talking points in Orlowski’s recent evaluation of Sweden’s approach to the disease (23). Similarly, “extensive local variation” in antibody levels has been used, in conjunction with classical (homogenous mixing) estimates of herd immunity, to argue that we are not close to reaching herd immunity (8). However, our model suggests that such variation is perfectly compatible, and, indeed, expected to be associated with, herd immunity.

**Table 1:** Results of Serological Testing for Antibodies in New York City, USA, and Lombardy, Italy, April 2020. More recent results from 1.46M antibody tests in New York City suggest an overall prevalence of 27% with values in various ZIP codes ranging from 12.6% to 51.6% (22). This real-world variation, at the ZIP code scale, mirrors the county-level variation predicted by our model.

| Location | Date | Prevalence | Reference |
| --- | --- | --- | --- |
| New York City, USA | April 2020 | 19.3% | (30) |
| Lombardy, Italy | April 2020 | 23.0% | (31) |

On the other hand, there are emerging glimpses of and discussion about pockets of immunity (24,25). Although they paid a terrible price, COVID-19 hospitalizations and deaths have recently fallen off dramatically in Manaus, Brazil, a one-time hot spot. There, serological testing for antibodies has never surpassed the 20% level, but some are hypothesizing that the strong peak and widespread community transmission created a protective level of herd immunity (25). Antibody testing has a number of known issues: (i) the quality of tests, (ii) the low sensitivity at early stages post-infection, (iii) the observed rapid falloff of antibodies (26), and (iv) the fact that antibody production appears to be strongly correlated with disease severity (and there are many mild cases). For these reasons, observed low antibody levels in seroprevalence studies and herd immunity may be compatible in some locales. As Figure 2 and Figure 3 illustrate, we should expect a substantial variation in case numbers across geographies with an overall threshold for herd immunity that is below the classical value.

Beyond our work and that of Britton, there are additional approaches to the herd immunity question. Gomes and collaborators (16,17) have explored variation in individuals. That is, they introduce models that incorporate variation in susceptibility of individuals in a much more systematic way than our binary classification. Their results, which suggest that herd immunity may be reached when as little as 25% of the population is infected, rely on assumptions about the coefficient of variation of the susceptibility. More information about this variation is needed to clarify the validity of their conclusions, but their work broadly supports the notion that heterogeneity lowers herd immunity thresholds.

Yet another approach, due to Di Lauro et al. (15), examines network heterogeneity in which interventions are modeled in two ways. When considered as changes in transmission rates, the analysis confirms the results described above, namely that in the presence of degree heterogeneity, the initial outbreak will grant herd immunity with significantly fewer infections than equivalent models with less or no degree heterogeneity. However, when the authors model interventions by modifying the contact network, then subtleties may arise. For example, if highly connected nodes are shielded, due to social distancing or other isolation strategies, from infection during an initial outbreak, upon reconnection, the secondary outbreak may in fact be more substantial than the first.

There are a number of limitations in this work. The model is, of course, an approximation, and the uniform, multiplicative imposition of nonpharmaceutical interventions (e.g., social distancing, use of face masks, etc.) across all geographies is artificial. Our implementation of partial immunity is also simplistic with some arbitrariness in the parameter choices. The mathematical model confers permanent immunity on recovered individuals, and no one knows how long actual immunity lasts. Reinfections are beginning to be reported and, among other possibilities, the virus could mutate. Thus, there are a number of reasons to be cautious. Social distancing and other mitigation strategies have been imposed in a highly nonuniform way (in time, space, implementation, compliance) across the nation. As these behaviors and strategies are relaxed, the possibility of exposing previously-shielded-but-central-to-transmission nodes and generating new outbreaks is very real.

Despite these limitations, the estimates presented here, in combination with other efforts (13– 17), have identified heterogeneity in contact structures (i.e., mixing between age groups or spatial distribution) and individual variation (in terms of infectivity, susceptibility, partial immunity, resistance) as key factors that reduce the disease-induced herd immunity level. It is likely that models featuring more realistic combinations of these factors would suggest even lower levels of disease-induced herd immunity. Natural next steps are to attempt to more precisely quantify the level of herd immunity for COVID-19 and to turn these insights to practical use. For example, while optimal vaccination strategies are much discussed in the literature (27), vaccine deployment is a complex issue entailing intersecting concerns from political, ethical, medical, production, and supply chain arenas. Recently released discussion guidelines from the National Academy of Medicine (28) focus on protection of individuals (e.g., critical workers including healthcare providers and emergency services personnel together with vulnerable populations). The calculations presented here suggest an alternative strategy; to limit future transmission and spread of the disease, we should consider not just *who* should be vaccinated, but *where* those vaccinations will do the most good.

## Data Availability

all data is available via the New York Times

https://github.com/nytimes/covid-19-data

## Notes

### Competing Interest Statement

The authors have declared no competing interest.

### Funding Statement

no external funding to report

### Author Declarations

This research is deemed exempt by the UnitedHealth Group IRB

## REFERENCES

1. Mills C, Robins J, Lipsitch M. Transmissibility of 1918 pandemic influenza. Nature. 2004;432(7019):904–906.

2. Johnson CY. A coronavirus vaccine won’t change the world right away. Washington Post. August 2, 2020.

3. DeMarco C. COVID-19 herd immunity: 7 questions, answered. MD Anderson Cancer Center. https://www.mdanderson.org/cancerwise/what-is-covid-19-coronavirus-herd-immunity-when-will-we-achieve-herd-immunity.h00-159383523.html.Published July 17, 2020. Accessed September 11, 2020.

4. Fontanet, A, Cauchemez, S. COVID-19 herd immunity: where are we? Nat Rev Immunol. 2020;20(10):583–584.

5. Randolph H, Barreiro L. Herd Immunity: Understanding COVID-19. Immunity. 2020;52(5):737–741.

6. Kwok K, Lai F, Wei W, Wong S, Tang J. Herd immunity –estimating the level required to halt the COVID-19 epidemics in affected countries. J Infect. 2020;80(6):e32–e33.

7. Altmann D, Douek D, Boyton R. What policy makers need to know about COVID-19 protective immunity. Lancet. 2020;395(10236):1527–1529.

8. Dey P, Bradley E, and Howard P. Don’t count on herd immunity for COVID-19 yet. A vaccine is the best way to get there. USA TODAY. August 18, 2020.

9. Wu J, Leung K, Leung G. Nowcasting and forecasting the potential domestic and international spread of the 2019-nCoV outbreak originating in Wuhan, China: a modelling study. Lancet. 2020;395(10225):689–697.

10. Taubenberger J, Morens D. 1918 Influenza: the mother of all pandemics. Emerg Infect Dis. 2006;12(1):15–22.

11. Fraser C, Donnelly C, Cauchemez S, et al. Pandemic Potential of a Strain of Influenza A (H1N1): Early Findings. Science. 2009;324(5934):1557–1561.

12. Centers for Disease Control and Prevention. 2009 H1N1 Pandemic. https://www.cdc.gov/flu/pandemic-resources/2009-h1n1-pandemic.html. Published June 11, 2019. Accessed May 31, 2020.

13. Britton T, Ball F, Trapman P. A mathematical model reveals the influence of population heterogeneity on herd immunity to SARS-CoV-2. Science. 2020;369(6505):846–849.

14. Lourenco J, Pinotti F, Thompson C, et al. The impact of host resistance on cumulative mortality and the threshold of herd immunity for SARS-CoV-2. medRxiv. 2020. 2020:2020.07.15.20154294. (doi:10.1101/2020.07.15.20154294). Accessed July 16, 2020.

15. Di Lauro F, Berthouze L, Dorey M, Miller J, Kiss I. The impact of network properties and mixing on control measures and disease-induced herd immunity in epidemic models: a mean-field model perspective. 2007.06975 [physics, q-bio]. Published online July 14, 2020. Accessed August 17, 2020.

16. Gomes M, Corder R, King J, et al. Individual variation in susceptibility or exposure to SARS-CoV-2 lowers the herd immunity threshold. medRxiv. 2020. (doi:10.1101/2020.04.27.20081893) Accessed July 16, 2020.

17. Aguas R, Corder R, King J, et al. Herd immunity thresholds for SARS-CoV-2 estimated from unfolding epidemics. medRxiv. 2020. (doi:10.1101/2020.07.23.20160762) Accessed July 16, 2020.

18. Grifoni A, Weiskopf D, Ramirez S, et al. Targets of T Cell Responses to SARS-CoV-2 Coronavirus in Humans with COVID-19 Disease and Unexposed Individuals. Cell. 2020;181(7):1489-1501.e15.

19. Meckiff B, Ramírez-Suástegui C, Fajardo V, et al. Single-cell transcriptomic analysis of SARS-CoV-2 reactive CD4+ T cells. bioRxiv. 2020. (doi:10.1101/2020.06.12.148916). Accessed August 18, 2020.

20. Braun J, Loyal L, Frentsch M, et al. SARS-CoV-2-reactive T cells in healthy donors and patients with COVID-19. Nature. 2020. (doi:10.1038/s41586-020-2598-9). Accessed July 29, 2020.

21. Verdecchia P, Cavallini C, Spanevello A, et al. The pivotal link between ACE2 deficiency and SARS-CoV-2 infection. Eur J of Intern Med. 2020;76:14–20.

22. Goldstein J. 1.5 Million Antibody Tests Show What Parts of N.Y.C. Were Hit Hardest - New York Times. August 19, 2020.

23. Orlowski E, Goldsmith D. Four months into the COVID-19 pandemic, Sweden’s prized herd immunity is nowhere in sight. J R Soc Med. 2020;113(8):292–298.

24. Mandavilli A. What if ‘Herd Immunity’ Is Closer Than Scientists Thought? New York Times. August x17, 2020.

25. McCoy T, Traiano H. In the Brazilian Amazon, a sharp drop in coronavirus sparks questions over collective immunity. Washington Post. August 24, 2020.

26. Ibarrondo F, Fulcher J, Goodman-Meza D, et al. Rapid Decay of Anti–SARS-CoV-2 Antibodies in Persons with Mild Covid-19. N Engl J Med. 2020;383(11):1085–1087.

27. Keeling M, Shattock A. Optimal but unequitable prophylactic distribution of vaccine. Epidemics. 2012;4(2):78–85.

28. National Academies. A Framework for Equitable Allocation of Vaccine for the Novel Coronavirus. https://www.nationalacademies.org/our-work/a-framework-for-equitable-allocation-of-vaccine-for-the-novel-coronavirus. Accessed September 15, 2020.

29. Nytimes/Covid-19-Data. New York Times; 2020. https://github.com/nytimes/covid-19-data. accessed September 17, 2020.

30. Stadlbauer D, Tan J, Jiang K, et al. Seroconversion of a city: Longitudinal monitoring of SARS-CoV-2 seroprevalence in New York City. medRxiv. 2020. (doi:10.1101/2020.06.28.20142190). Accessed July 16, 2020.

31. Percivalle E, Cambiè G, Cassaniti I, et al. Prevalence of SARS-CoV-2 specific neutralising antibodies in blood donors from the Lodi Red Zone in Lombardy, Italy, as at 06 April 2020. Euro Surveill. 2020;25(24):2001031.

