## Supplemental Materials for "Updating Herd Immunity Models for the U.S. in 2020: Implications for the COVID-19 Response"

### 1. Network model

The model aims to combine both the ease of parametrization associated with SIR-type models while also leveraging features of networks to provide realistic pathways for disease transmission at a state and regional level. The model is structured as follows:

- The backbone of the model is a network of nodes and edges. Each node represents a US county (or county equivalent), and the edges connecting nodes are derived from transit data. In particular, short-range edges are computed from commuter data while long-range edges are computed from actual flight data.
- Dynamics within each node are principally driven by classical SIR-type dynamical models from mathematical epidemiology. These within-county models are weakly coupled to their neighbors on the graph. That is, a susceptible individual in county $k$ is assumed to randomly mix with the population of county $k$ and, in addition, this susceptible individual has a small chance of encountering an infectious individual from a neighboring county. Thus, an outbreak initially localized in a single county may, depending on parameter settings, spread to neighboring counties or, given the presence of a long-range air travel link, jump across the country.

As the foundation of our model, we take a four-compartment model with compartments for susceptible, infectious, recovered, and deceased individuals; there is evidence that these simple models describe the outbreaks in China and Italy well [3]. We ignore vital dynamics. Thus,

$$S, I, R, D \in [0, \infty)$$

represent the number of susceptible, infectious, recovered, and deceased people in a fixed population of size $P$ ($S + I + R + D = P$). Then, the model takes the form

$$S' = -\beta SI/P\,, \tag{1.1a}$$

$$I' = \beta SI/P - (\gamma + \delta)I\,, \tag{1.1b}$$

$$R' = \gamma I\,, \tag{1.1c}$$

$$D' = \delta I\,. \tag{1.1d}$$

Note that $'$ denotes differentiation with respect to time ($t$, in days) and

$$(S + I + R + D)' = 0\,,$$

whence $S + I + R = P$ where $P$ is the (constant) total population. In (1.1), constant parameters $\beta$, $\gamma$, and $\delta$ are positive and represent the *contact/infection rate*, the relative *mean removal rate*, and the *disease-induced relative mortality rate*, respectively. In this case,

- $\beta$ has the units of $\frac{1}{\text{days}}$ and represents a variety of intertwined phenomena that relate to the transmission of disease,
- $\gamma$ has the same units and is the relative rate of disease progression, and
- $\delta$ is the disease-induced relative death rate for infectious people.

*Date*: September 17, 2020.

For model (1.1), the *reproduction number* [6, 5] is the dimensionless quantity:

$$R_0 = \frac{\beta}{\gamma + \delta}\,. \tag{1.2}$$

As is well known, we require $R_0 < 1$ to eliminate the disease. For model (1.1), it is straightforward to verify that that $R_0 < 1$ implies that the disease-free equilibrium $(S_0, 0, 0, 0)$ is locally asymptotically stable.

1.1. **Abstract formulation.** We describe a phenomenological model for network spreading of disease using the system (1.1) as the basis for our disease model. We start with a family of graphs, $G^{(\ell)} = (V, E^{(\ell)})$, $\ell = 1, 2, \dots, M$ on a common set of $N$ vertices (nodes):

$$V = \{v_1, v_2, \dots, v_N\}\,.$$

Here, the nodes represent a fixed set of geographic locations, and the various sets of edges, $E^{(\ell)}$ represent different connections between these geographies. By an abuse of nomenclature, we will refer to this collection of graphs on a common set of nodes as "the network." We denote by $S_k, I_k, R_k, D_k$ the susceptible, infectious, recovered, and deceased people at node $k$. Similarly, we denote the population of vertex $k$ by $P_k$, whence

$$P = \sum_{k=1}^{N} P_k$$

is the total population in the model, and we write $\boldsymbol{P} \in \mathbb{R}^N$ for the vector of populations at each nodes.

We denote by $\mathsf{A}^{(\ell)} = [a_{kj}^{(\ell)}]$ the adjacency matrix for the graph $G^{(\ell)}$. That is,

$$a_{kj}^{(\ell)} = \begin{cases} 0 & \text{if} \quad (k,j) \notin E^{(\ell)}\,, \\ 1 & \text{if} \quad (k,j) \in E^{(\ell)}\,. \end{cases} \tag{1.3}$$

We don't allow self-connections. Each adjacency matrix $\mathsf{A}^{(\ell)}$ is thus symmetric with zeros on the diagonal. We consider the $k$th node in the graph, and we assume that the infection rate at that node is proportional to mixing in that node and is also weakly coupled to the number of infected people in neighboring nodes:

$$I_k' = \frac{\beta_k S_k}{P_k}\left[ I_k + \sum_{\ell=1}^{M} \sigma^{(\ell)} \sum_{j=1}^{N} a_{kj}^{(\ell)} I_j \right] - (\gamma + \delta) I_k\,. \tag{1.4}$$

Here, each $\sigma^{(\ell)} \geq 0$ is a small parameter measuring the strength of the coupling to the neighboring nodes in the $\ell$th graph that makes up the network. Evidently, $\sigma^{(\ell)} = 0$ means no coupling. Conversely, a value of $\sigma^{(\ell)} = 1$ means a susceptible person at node $k$ is equally likely to interact with an infected person at node $k$ as any other neighboring node. If $\sigma > 1$, infections at node $k$ would be generated more from nodes outside of node $k$ than within of node $k$.

To calculate $R_0$, the reproduction number for the network model, we find the largest eigenvalue of the matrix whose $(j,k)$ element is $\frac{\beta_k}{\gamma+\delta} \sum_{\ell=1}^{M} \sigma^{(\ell)} a_{kj}^{(\ell)}$.

1.2. **Building realistic networks.** In practice, we now consider two edge sets on the same set of nodes. And, for nodes, we select a $\mathsf{A}^{\text{short}} = [a_{\ell m}^{\text{short}}]$ the *short range* adjacency matrix representing geographic proximity, and suppose that or $\mathsf{A}^{\text{long}} = [a_{\ell m}^{\text{long}}]$ represents long-range (e.g., air link) connections. Each matrix is thus symmetric with zeros on the diagonal.

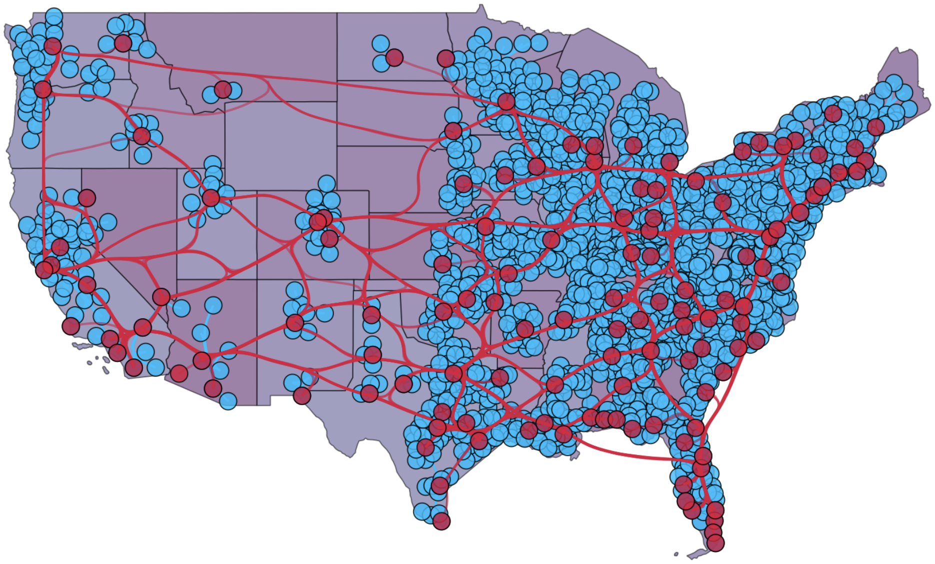

FIGURE 1. The network built for our model.

We consider the $k$th node in the graph, and we assume that the infection rate at that node is proportional to mixing in that node and is also weakly coupled to the fraction of infected people in neighboring nodes both near and far:

$$S_k' = -\frac{\beta S_k}{P_k}\left[I_k + \sigma_{\text{short}}\sum_{j=1}^{N} a_{kj}^{\text{short}} I_j + \sigma_{\text{long}}\sum_{j=1}^{N} a_{kj}^{\text{long}} I_j\right], \tag{1.5a}$$

$$I_k' = \frac{\beta S_k}{P_k}\left[I_k + \sigma_{\text{short}}\sum_{j=1}^{N} a_{kj}^{\text{short}} I_j + \sigma_{\text{long}}\sum_{j=1}^{N} a_{kj}^{\text{long}} I_j\right] - (\gamma+\delta) I_k\,, \tag{1.5b}$$

$$R_k' = \gamma I_k\,, \tag{1.5c}$$

$$D_k' = \delta I_k\,, \tag{1.5d}$$

(1.5e)

Here, $\sigma_{\text{short}}, \sigma_{\text{long}} \geq 0$ are small dimensionless parameters measuring the strength of the coupling to the neighboring proximate and far-away nodes, respectively.

1.2.1. *Data, filters, and calibration.* To construct the graph $\mathsf{A}^{\text{long}}$ and $\mathsf{A}^{\text{short}}$ we allow edges if at least 1,000 people travel on each edge each day. This data is publicly available from the US Bureau of Transportation Statistics.

In order to fit the model we minimize the difference between model predicted deaths and deaths as reported by the New York Times This gives a value of $\beta_k$ at each node as well as values for $\sigma_{\text{short}}$ and $\sigma_{\text{long}}$. We only attempt these fittings in the case of counties with at least 33 deaths. As of August 25, 2020 this was 544 of the 3,1829 counties considered in our graph reach this threshold.

For $\gamma$ and $\delta$ we select values consistent with the literature [1].

1.3. **Initial Conditions.** In [2] they assume that $s_j(0) = 1$ for all $j$ in order to simplify. The effect on the final value of herd immunity is small but nontrivial. To explore this dependence we present results in Figure 2 for herd immunity levels as presented by Britton $(1 - s_j(\infty))$, as calculated by assuming .25% of the population is infected at every node at time 0, and as calculated by assuming

2.5% of the population is infected at every node at time 0. In the main body, we report the version assuming .25% of the population is initially infected.

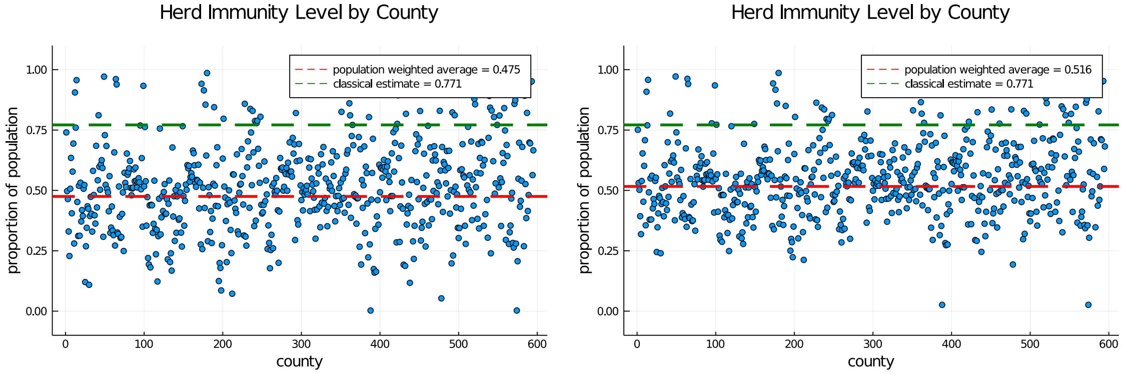


FIGURE 2. Herd immunity levels calculated as in [2], with $i_j(0) = .0025$ and $i_j(0) = .0250$

### 2. CROSS-REACTIVITY

To model cross-reactivity we follow [4], extending it to our network model, and assume the infectious population is broken into two compartments, those who are infectious but have cross reactive T-cells ($I_k^{(c)}$) and those who do not ($I_k^{(n)}$). Thus, (1.5) are replaced by

$$S_k' = -\frac{\beta S_k}{P_k}\left[I_k^{(n)} + \mu I_k^{(c)} + \sigma_{\text{short}}\sum_{j=1}^{N} a_{kj}^{\text{short}}(I_j^{(n)} + \mu I_j^{(c)}) + \sigma_{\text{long}}\sum_{j=1}^{N} a_{kj}^{\text{long}}(I_j^{(n)} + \mu I_j^{(c)})\right], \tag{2.1a}$$

$$I_k^{(c)\prime} = f_0\frac{\beta S_k}{P_k}\left[I_k^{(n)} + \mu I_k^{(c)} + \sigma_{\text{short}}\sum_{j=1}^{N} a_{kj}^{\text{short}}(I_j^{(n)} + \mu I_j^{(c)}) + \sigma_{\text{long}}\sum_{j=1}^{N} a_{kj}^{\text{long}}(I_j^{(n)} + \mu I_j^{(c)})\right] - (\gamma_c + \delta_c) I_k^{(c)}, \tag{2.1b}$$

$$I_k^{(n)\prime} = (1 - f_0)\frac{\beta S_k}{P_k}\left[I_k^{(n)} + \mu I_k^{(c)} + \sigma_{\text{short}}\sum_{j=1}^{N} a_{kj}^{\text{short}}(I_j^{(n)} + \mu I_j^{(c)}) + \sigma_{\text{long}}\sum_{j=1}^{N} a_{kj}^{\text{long}}(I_j^{(n)} + \mu I_j^{(c)})\right] - (\gamma + \delta) I_k^{(n)}, \tag{2.1c}$$

$$R_k' = \gamma I_k^{(n)} + \gamma_c I_j^{(c)}), \tag{2.1d}$$

$$D_k' = \delta I_k^{(n)} + \delta_c I_j^{(c)}, \tag{2.1e}$$

(2.1f)

where $f_0$ denotes the fraction of infected individuals who are have cross-reactive T-cells, $1/\gamma_c$ is the mean infectious period for a person who has cross-reactive T-cells, $\delta_c$ is the mean mortality rate for cross-reactive individuals, and having cross reactive T-cells reduces infectiousness by a factor of $\mu$.

In what follows, we vary $f_0$ and choose $\gamma_c$ and $\delta_c$ such that the infectious period is shortened by 1 day and the mortality rate is reduced by 10% for those with cross-reactive T-cells. We choose $\mu = .85$. In addition, we use seed the model with .25% of the population infected (split evenly between reactive and non-reactive).

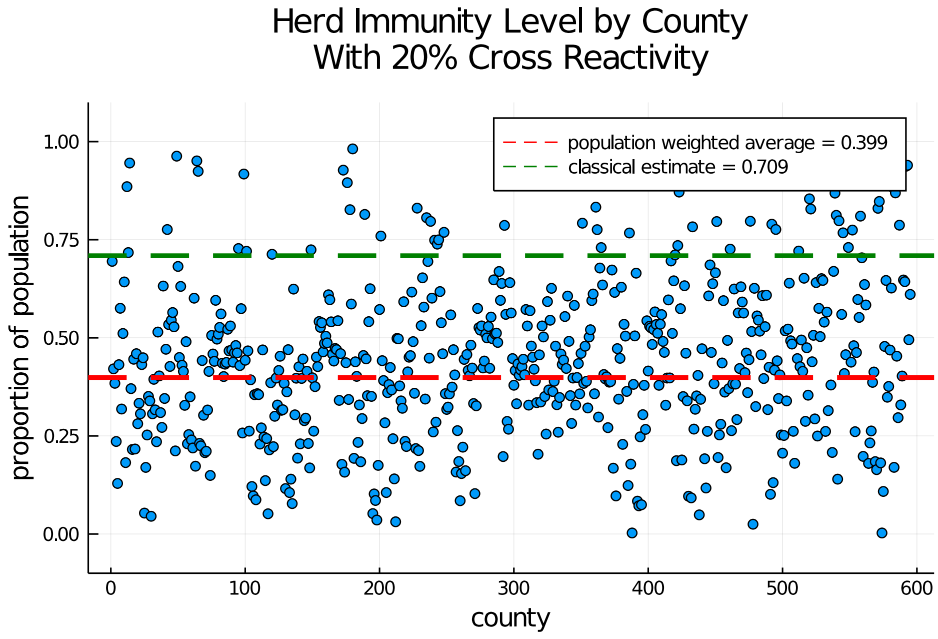


FIGURE 3. Herd immunity levels when we assume 20% of the population has cross-reactive T-cells

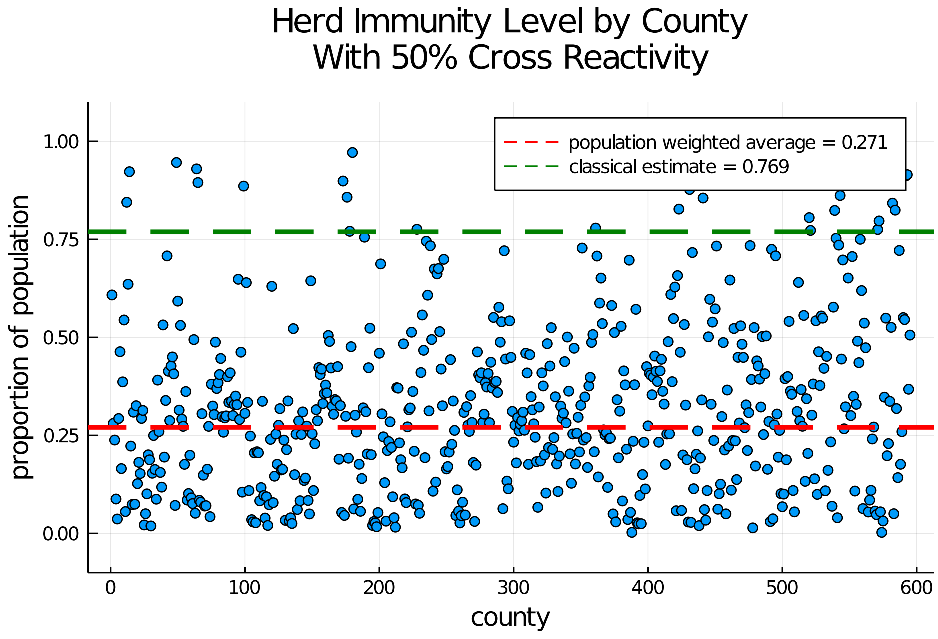


FIGURE 4. Herd immunity levels when we assume 50% of the population has cross-reactive T-cells

### REFERENCES


[1] Yinon M Bar-On, Avi Flamholz, Rob Phillips, and Ron Milo. SARS-CoV-2 (COVID-19) by the numbers. *eLife*, 9:e57309, March 2020.

[2] Tom Britton, Frank Ball, and Pieter Trapman. A mathematical model reveals the influence of population heterogeneity on herd immunity to SARS-CoV-2. *Science*, June 2020.

[3] Diego Caccavo. Chinese and Italian COVID-19 outbreaks can be correctly described by a modified SIRD model. *medRxiv*, 2020.

[4] Zhilan Feng, Sherry Towers, and Yiding Yang. Modeling the effects of vaccination and treatment on pandemic influenza. *The AAPS journal*, 13(3):427–437, 2011.

[5] Matt Reynolds. What is the coronavirus R number and why is it important? *Wired UK*, May 2020.

[6] Pauline van den Driessche. Reproduction numbers of infectious disease models. *Infectious Disease Modelling*, 2(3):288–303, June 2017.
